# Revised spectral metrics for body surface measurements of gastric electrophysiology

**DOI:** 10.1101/2022.07.05.22277284

**Authors:** Gabriel Schamberg, Chris Varghese, Stefan Calder, Stephen Waite, Jonathan Erickson, Greg O’Grady, Armen A. Gharibans

## Abstract

**Background:** Electrogastrography (EGG) non-invasively evaluates gastric function but has not achieved common clinical adoption due to several technical limitations. Body Surface Gastric Mapping (BSGM) has been introduced to overcome these limitations, but pitfalls in traditional metrics used to analyze spectral data remain unaddressed. This study critically evaluates five traditional EGG metrics and introduces improved BSGM spectral metrics, with validation in a large cohort.

**Methods:** Pitfalls in five EGG metrics were assessed (dominant frequency, percentage time normogastria, amplitude, power ratio, and instability coefficient), leading to four revised BSGM spectral metrics. Traditional and revised metrics were compared to validate performance using a 100 BSGM subject database (30 min baseline; 4-hrs postprandial), recorded using Gastric Alimetry (Alimetry, New Zealand).

**Key Results:** BMI and amplitude were highly correlated (r=-0.57, p<0.001). We applied a conservative BMI correction to obtain a *BMI-adjusted amplitude* metric (r=-0.21, p=0.037). Instability coefficient was highly correlated with both dominant frequency (r=-0.44, p<0.001), and percent bradygastria (r=0.85, p<0.001), in part due to conflation of low frequency transients with gastric activity. This was corrected by introducing distinct gastric frequency and stability metrics (*Principal Gastric Frequency* and *Gastric Alimetry Rhythm Index (GA-RI))* that were uncorrelated (r=0.14, p=0.314). Only 28% of subjects showed a maximal averaged amplitude within the first postprandial hour. Calculating *Fed:Fasted Amplitude Ratio* over a 4-hr postprandial window yielded a median increase of 0.31 (IQR 0-0.64) above the traditional ratio.

**Conclusions & Inferences:** The revised metrics resolve critical pitfalls impairing the performance of traditional EGG, and should be applied in future BSGM spectral analyses.

## Introduction

Chronic gastroduodenal symptoms affect >8% of the population and impart a substantial quality of life and economic burden.^1,2^ Existing diagnostic approaches for assessing the contribution of gastric dysmotility are controversial, and new tests are needed to aid diagnosis.^3,4^

Historically, gastric electrophysiological abnormalities were clinically assessed using electrogastrography (EGG) with sparse epigastric electrodes. Although EGG identified group-level differences between patients and controls in multiple adult and pediatric disorders,^5–8^ it did not achieve common clinical adoption due to several technical limitations, including inability to account for gastric anatomical variability, poor spatial resolution, and pitfalls in handling artifacts.^9–12^

Body surface gastric mapping (BSGM) overcomes these limitations, with recent studies showing capability to define novel disease subgroups with symptom correlations.^9,13,14^ BSGM is performed using a recently-developed medical device (Gastric Alimetry®, Alimetry, New Zealand) comprising a conformable high-resolution array, wearable Reader, and validated symptom-logging App.^13,15,16^ Using Gastric Alimetry, we have compiled a large database of high-quality recordings encompassing 4.5 hours (30 minutes preprandial, 4 hours postprandial).^13,16^ Compiling this database enabled us to critically re-evaluate spectral metrics previously applied in EGG, generating fresh insights into additional pitfalls that contributed to EGG’s failure to become a routine diagnostic tool.

In this study, we address these pitfalls and introduce revised spectral metrics for BSGM, demonstrating utility in a large dataset. While BSGM provides both spectral metrics and novel spatial metrics (wave patterns), ^13,16,17^ the current analysis focuses on spectral analytics.

## Methods

Traditional EGG metrics are concerned with frequency, power (or amplitude), and variability, derived from spectrograms calculated using a fast Fourier transform with a sliding window.^18^ **Supplementary Table 1** defines five key EGG metrics: dominant frequency, percentage normal / tachygastria / bradygastria, amplitude, power ratio, and instability coefficient of the dominant frequency.

The pitfalls of each metric are discussed below together with the justification of revised metrics, which are discussed in greater detail in **Supplementary Table 2**.

### Pitfall 1: Effect of BMI on Amplitude

Adipose tissue reduces extracellular potential transmission; hence higher BMIs attenuate EGG amplitudes.^19^ While this relationship is known, it has not been robustly quantified, preventing amplitude from being compared across subjects or studies. To resolve this issue, we introduce *BMI-adjusted amplitude*, employing a multiplicative regression model to correct amplitude attenuation.

### Pitfall 2: Conflation of low frequency transients with stable gastric activity

Dominant frequency is typically calculated as the frequency associated with the highest power in the spectrum for a given period/window. This approach can be problematic, because normal cutaneous gastric myoelectrical recordings commonly show transient high-amplitude bursts of low frequency noise (<2 cpm) occurring concurrently with stable gastric activity of normal frequency,^12^ likely representing colonic activity,^20,21^ or artifacts.^10,12^ Dominant frequency calculations may therefore conflate non-gastric low frequency transients with stable gastric activity. Note that while patients with gastric neuromuscular dysfunction also show substantial spectral instability within this same frequency range, this typically occurs when coordinated gastric activity is intermittent or absent.^16^

To resolve this issue, we developed robustly distinct frequency and stability metrics. *Principal Gastric Frequency* identifies only the sustained frequency associated with the most stable spectral oscillations, as determined by the separate stability metric, *Gastric Alimetry Rhythm Index (GA-RI)* (discussed below).

### Pitfall 3: Misuse of coefficient of variation

The formula for the instability coefficient (coefficient of variation) serves as a normalized measure of dispersion. However, this metric is inappropriate for gastric rhythm because it implicitly and incorrectly assumes that a dominant frequency varying around a lower frequency is less stable than one varying around a higher frequency (see **Supplementary Table 2**).

To resolve this issue, we introduce *Gastric Alimetry Rhythm Index (GA-RI)™*, a proprietary measure of rhythmic stability quantifying the extent to which activity is concentrated within a narrow frequency band over time relative to the residual spectrum. We observed that BMI confounds stability metrics (see Results), hence *GA-RI* also includes a BMI adjustment.

### Pitfall 4: Variability in meal response timing

EGG power normally increases after a meal,^13,22^ however, we observe substantial variability in the timing of the onset and duration of this response.^13,16,23^ Therefore, if meal response metrics employ only a brief fixed window (e.g. 45 minute recordings), meal responses may be underestimated.

We therefore introduce *Fed:Fasted Amplitude Ratio*, a ratio of the maximum amplitude in any single 1-hour over a 4-hr postprandial period, to the preprandial period.

### Experimental Methods

The relative performance of the revised metrics was evaluated in a large existing BSGM database (100 participants; 4.5-hr recordings), comprising healthy controls. For full experimental methodology, refer.^13,16,24^ Ethical approval was granted by the Auckland Health Research Ethics Committee (AH1130) and all subjects provided informed consent.

## Results

### BMI-Adjusted Amplitude

Regression analyses showed significant associations between BMI and average amplitude (r=-0.57, p<0.001; **Fig. 1A**). **Fig. 1B**. reports the relationship between BMI and the revised *BMI-adjusted amplitude* metric. A conservative adjustment was achieved with a small residual effect (r=-0.21, p=0.037), intentionally designed to avoid over-adjustment in excessively high BMIs.

**Figure 1.**
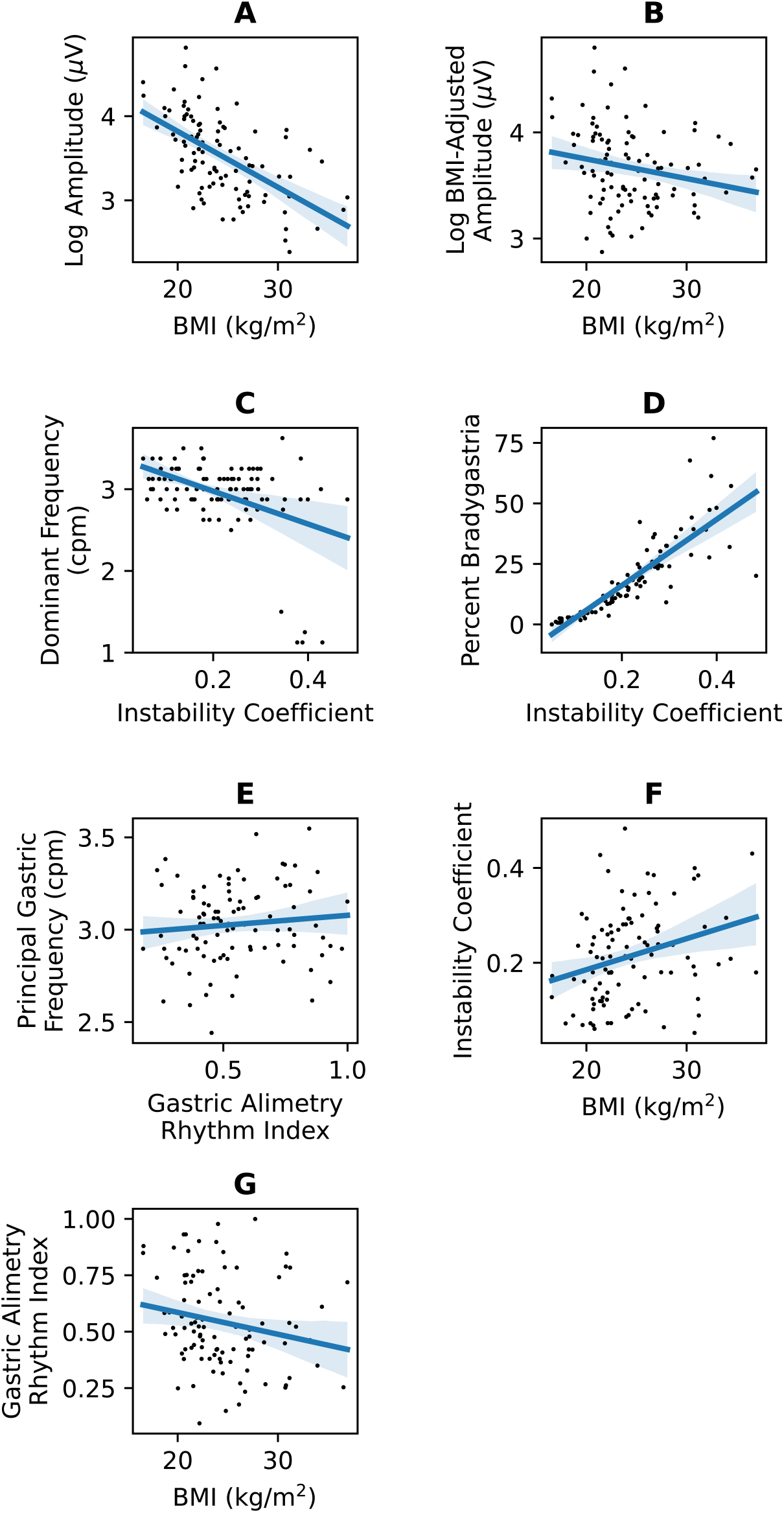
Relevant associations between traditional EGG metrics, revised BSGM metrics, and/or patient characteristics. (**A**) BMI and log average amplitude (**B**) BMI and log BMI-Adjusted Amplitude (**C**) Instability coefficient and dominant frequency (**D**) Instability coefficient and percent bradygastria (**E**) Gastric Alimetry Rhythm Index and Principal Gastric Frequency (**F**) BMI and instability coefficient (**G**) BMI and Gastric Alimetry Rhythm Index

### Principal Gastric Frequency and GA-RI

Dominant frequency was negatively correlated with instability coefficient (r=-0.44, p<0.001; **Fig. 1C**), with incorrect identification of dominant frequency as <1.5 cpm contributing in a subset due to misclassification of low frequency transients as bradygastric activity. **Fig. 1D** reveals a strong correlation between instability coefficient and percent bradygastria (r=0.85, p<0.001); therefore, the extent to which percent bradygastria indicates abnormally slow but coordinated gastric activity, as opposed to high variability in dominant frequency, is unclear.

By contrast, the ability of the revised *GA-RI* and *Principal Gastric Frequency* metrics to independently capture frequency and stability is reported in **Fig. 1E** (r=0.10, p=0.314).

The relationship between BMI and instability coefficient is shown in **Fig. 1F** (r=0.28, p=0.004). As such, the GA-RI metric includes a conservative BMI adjustment (**Fig. 1G**; r=-0.20, p=0.044).

**Fig. 2A** shows an example problematic control spectrogram with both sustained activity at a normal frequency and a large volume of transient low frequency noise at <2 cpm. Here, the traditional dominant frequency metric identifies the gastric frequency as 1.1 cpm, whereas the revised *Principal Gastric Frequency* metric identifies the gastric frequency as 2.9 cpm. The *Principal Gastric Frequency* metric therefore correctly indicates that this subject has a normal frequency, while the low frequency activity is separately captured by assigning a *GA-RI* of 0.18, indicating low background spectral stability.

**Figure 2.**
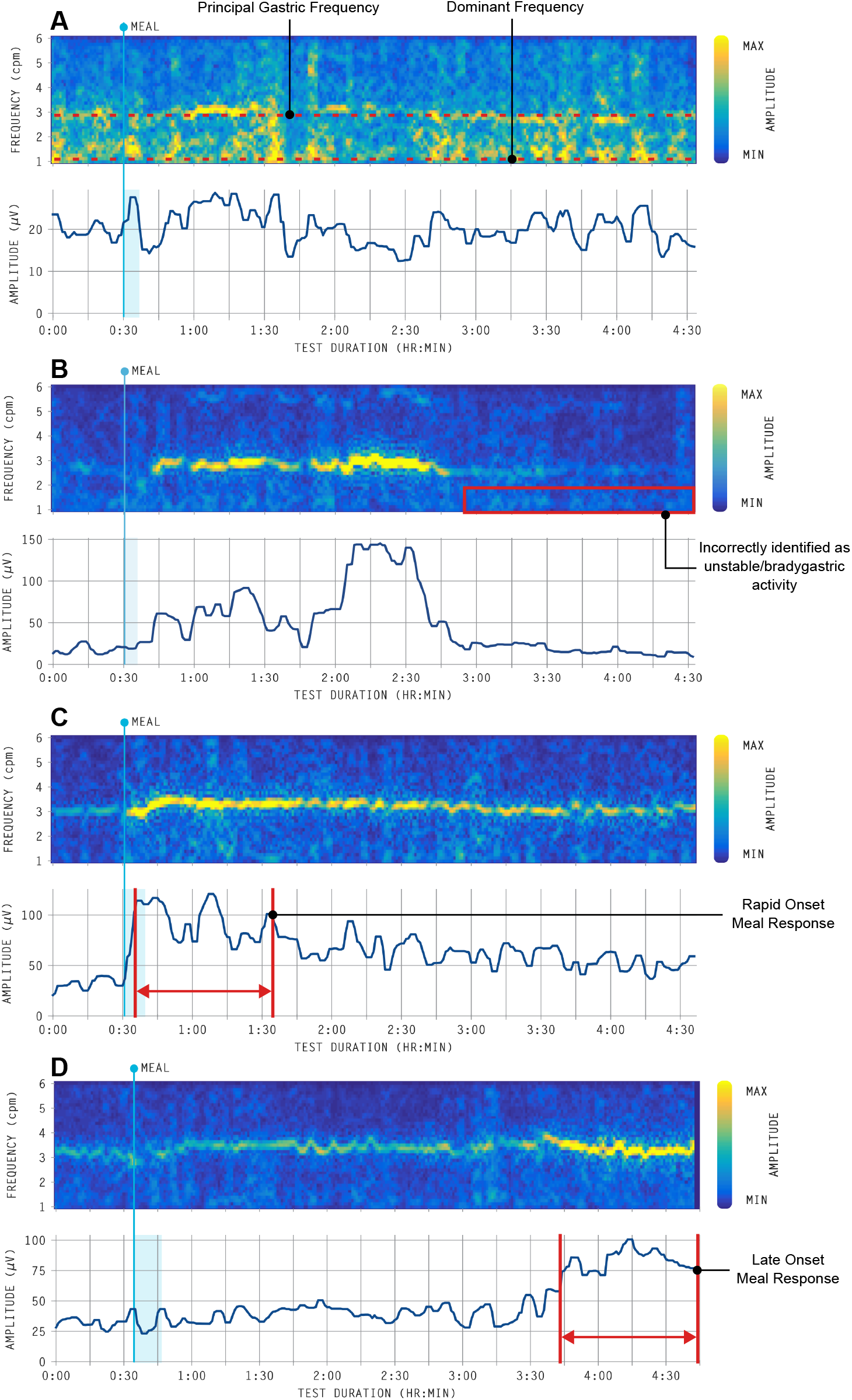
Representative spectrograms from healthy control subjects, recorded using the Gastric Alimetry system, for (**A**) Transient low frequency activity resulting in misclassified Dominant Frequency. (**B**) Inactive periods misclassified as unstable/bradygastric by traditional EGG metrics. (**C, D**) Examples of early vs late onset meal responses respectively; in these cases, using the entire postprandial average amplitude/power captures significant portions of relative inactivity, whereas a fixed window average (i.e., <1-hour postprandial) would miss either the early or delayed response.

*GA-RI* also improved on the problem of percent bradygastria and instability coefficient metrics being prone to indicating abnormal gastric activity during times where the stomach was simply inactive. An example is shown in **Fig. 2B**, depicting a spectrogram with normal periods of relative inactivity preprandially, and after the meal response. In this case, the GA-RI is 0.98 (indicating highly stable rhythmic activity), whereas percent bradygastria and instability coefficient metrics were 25% and 0.23 (above the median instability coefficient), respectively.

### Meal Response

Across the whole dataset, median peak amplitude occurred 1.6 hours postprandially (IQR 0.7-2.6). Only 28% of healthy controls had a maximal average amplitude in the first postprandial hour. When moving from a fixed amplitude ratio using the first hour postprandially to the *Fed:Fasted Amplitude Ratio* over a 4-hr period, the median increase was 0.31 (IQR 0-0.64). **Figs. 2C** and **2D** show examples of spectrograms with early and delayed-onset meal responses.

## Discussion

Despite extensive research, EGG did not achieve common clinical adoption owing to technical limitations that impacted clinical utility.^9,11^ BSGM using Gastric Alimetry has overcome many of these limitations,^13,14,16^ but pitfalls in traditional metrics have remained unaddressed. Using an extensive BSGM dataset, we critically evaluated past spectral metrics and present a revised metric set that resolves these pitfalls.

The revised metrics, together with the other technical and processing advances of BSGM,^10,13,15^ offer a substantially more rigorous foundation for advancing the clinical utility of cutaneous gastric electrophysiology, and will be incorporated into the Gastric Alimetry test. However, their clinical application also requires a comprehensive reference range, which is currently being compiled.^24^

Concerns regarding ‘percent bradygastria’ were addressed here, but problems with the interpretation of ‘percent tachygastria’ also warrant discussion. While many EGG studies report frequencies in the tachygastric range (e.g. >4 cpm),^5–8^ direct recordings and pacing studies show that human gastric frequencies rarely exceed ∼4.5 cpm.^25^ Higher frequencies yielded by EGG could therefore also potentially reflect non-gastric sources.^26^ Gastric Alimetry employs a tight spectral window of 1-6 cpm, capturing the vast majority of relevant gastric activity, while eliminating physiologically implausible data <1 cpm and >6 cpm.^12,26^

Our study also revealed the importance of correcting stability metrics for BMI. Previous studies have found relationships between obesity and increased percentage of EGG activity beyond the normal frequency range,^19^ however such data should be interpreted cautiously without BMI-correction.

Significant post-processing research remains before the full potential of BSGM is realized. Spatial analytics encompassing wave patterns have demonstrated clear potential to further improve diagnostic yield, but still require refinement, optimization and standardization.^13,14,16,17^ In addition, analyses of the shape and dynamics of postprandial BSGM amplitude curves could offer new insights into gastric dysfunction.^13^ Nevertheless, spectral metrics alone offer fundamental value in characterizing disease states,^16,23^ and revising these to avoid pitfalls constitutes one more step toward realizing the clinical potential of non-invasive gastric diagnostics.

## Data Availability

Data are available upon reasonable request at the discretion of the corresponding author.

---

**Supplementary Table 1.** Definitions of five key traditional EGG metrics.^27–29^

| Metric | Definition |
| --- | --- |
| *Dominant Frequency* | The frequency (cycles per minute; cpm) associated with the strongest power in the overall spectrum of a recorded EGG signal per time point. A range of 2.6-3.7 cpm has been proposed based on healthy control studies, however, other ranges as broad as 2-4 cpm have also been applied.<sup>12,30</sup> |
| *Percentage Normal / Bradygastria / Tachygastria* | The percentage of time windows where the frequency associated with the highest power is within a predefined frequency range. This range has not been defined in a standardized or widely accepted manner, with 2-4 cpm being one commonly applied range (bradygastria <2 cpm; tachygastria >4 cpm), and >70% time in 'normogastria' typically considered normal.<sup>30,31</sup> |
| *Amplitude / Power* | The amplitude/power (μV/dB) associated with the dominant frequency in the overall spectrum, averaged over time. |
| *Amplitude / Power Ratio* | The ratio of the amplitude/power in the postprandial window to the amplitude/power in the preprandial window. |
| *Instability Coefficient of the Dominant Frequency* | The standard deviation of the dominant frequency divided by the mean of the dominant frequency over time. |

**Supplementary Table 2.** Revised BSGM Spectral Metrics

| Metric | Definition and Rationale |
| --- | --- |
| *BMI-Adjusted Amplitude* | The amplitude/power (μV/dB) associated with dominant frequency in the overall spectrum is dependent on BMI. We therefore implemented a conservative BMI-adjusted amplitude using a multiplicative regression. |
| *Principal Gastric Frequency* | Cutaneous gastric myoelectrical recordings in controls commonly show transient high-amplitude bursts of low frequency noise (<2 cpm), occurring concurrently with stable gastric activity of normal frequency.<sup>12</sup> Normal gastric activity is globally entrained to frequencies near to 3 cpm,<sup>32,33</sup> hence these transient bursts likely reflect colonic activity,<sup>20,34</sup> or artifacts.<sup>10,12</sup><br>Dominant frequency metrics define the highest average power across the spectra and may therefore conflate low-frequency high-amplitude transients with gastric activity.<sup>12</sup><br>The revised 'principal gastric frequency' metric instead identifies only the frequency associated with the most stable oscillations, as measured by a distinct new stability metric (GA-RI; see |
|  | below), distinctly separating measures of gastric frequency and stability. The principal gastric frequency metric therefore detects the intrinsic gastric frequency, as opposed to simply calculating the frequency with the highest average power including all spectral contents whether gastric in origin or otherwise.<br>Note that in contrast to controls, patients with gastric neuromuscular dysfunction and suspected ICC loss show substantial spectral instability within this same frequency range (due to summation of discoordinated wavefronts),<sup>35,36</sup> however this typically occurs when concurrent coordinated gastric activity is intermittent or absent.<sup>16</sup> |
| *Gastric Alimetry Rhythm Index (GA-RI)* | The formula for the instability coefficient is known as the coefficient of variation and serves as a normalized measure of dispersion (see **Supplementary Table 1**). While the coefficient of variation is used widely across statistical disciplines, its use as a measure of instability for the dominant gastric frequency poses a pitfall because it is frequency dependent.<br>For example, the coefficient of variation is correctly used in economics to assess investment risk (historical return divided by mean return), where a higher mean return would be a more stable investment (i.e., lower coefficient of variation). However, the use of this metric as a measure of instability for the dominant frequency falsely implies that a dominant frequency varying between a lower frequency (e.g., 2.4-2.6 cpm) would be less stable than one varying between a higher frequency (e.g., 3.4-3.6 cpm). In other words, instability coefficient metrics are intrinsically lower at higher dominant frequencies (**Fig. 1A**) despite there being no clinical or physiological justification for such a relationship.<br>To overcome this problem, we introduce the ‘Gastric Alimetry Rhythm Index’ (GA-RI), providing a measure of stability of the rhythmic gastric activity, by quantifying the extent to which activity is concentrated within a narrow frequency band over time relative to the residual spectrum. This proprietary metric improves on previous stability metrics in that it has no inherent dependence on the dominant frequency. As a result, the GA-RI enables independent assessment of the frequency and stability of gastric activity, i.e., it reflects the physiological evidence that it is possible to have abnormally high or low frequency gastric activity that is stable, or to have activity at a normal gastric frequency that is unstable.<sup>16,35,37</sup><br>Furthermore, the GA-RI includes a conservative BMI adjustment to account for the effect that signal attenuation has on the perceived relative strength of the gastric activity. |
| *Amplitude / Power Ratio* | Amplitude increase following a meal stimulus is a characteristic of healthy gastric function.<sup>13,22</sup> However, on visual inspection of 4-hr postprandial recordings, we have shown substantial variability in the timing of the onset and duration of the meal response.<sup>13,16,23</sup> |
|  | As a result, metrics based on an amplitude/power ratio, computing the relative postprandial to preprandial averaged amplitude/power using either the entire postprandial period or a fixed window within the postprandial period, are at risk of missing the meal response or underestimating the gastric response due to inclusion of low-amplitude activity within the selected postprandial window. In EGG, for example, meal response metrics have often been formulated based on the first 45 minutes of the postprandial period alone. This pitfall may not apply if water load stimuli are employed.<br>Reliably quantifying a meal response on an individual basis requires a metric that is robust to the inherent variability in meal response dynamics.<br>We therefore introduce the *Fed:Fasted Amplitude Ratio*, instead quantifying the observed meal response by taking a ratio of the maximum amplitude in any single 1-hour postprandial period to the amplitude in the preprandial period. Given that the goal of amplitude/power ratio metrics is to identify an increase in amplitude/power following meal consumption, it is important to have a metric that can quantify this increase across a cohort of subjects with significant natural variation in the timing of the meal response.<sup>13</sup><br>At the population level, meal responses would be significantly underestimated by a test including only a 1-hour or less of postprandial recordings. As with the other revised metrics, this metric is designed to capture a single precise characteristic of the gastric activity. Other characteristics of the meal response, such as the duration or time of onset, could be captured by additional targeted metrics in future should these characteristics be deemed physiologically or clinically relevant. |

## References

1. Sperber, A. D. et al. Worldwide Prevalence and Burden of Functional Gastrointestinal Disorders, Results of Rome Foundation Global Study. Gastroenterology 160, 99-114.e3 (2021).

2. Lacy, B. E., Weiser, K. T., Kennedy, A. T., Crowell, M. D. & Talley, N. J. Functional dyspepsia: the economic impact to patients. Aliment. Pharmacol. Ther. 38, 170–177 (2013).

3. Pasricha, P. J. et al. Progress in gastroparesis-a narrative review of the work of the Gastroparesis Clinical Research Consortium. Clinical Gastroenterology and Hepatology (2022) doi:10.1016/j.cgh.2022.05.022.

4. Pasricha, P. J. et al. Functional Dyspepsia and Gastroparesis in Tertiary Care are Interchangeable Syndromes With Common Clinical and Pathologic Features. Gastroenterology 160, 2006–2017 (2021).

5. Bhat, S. et al. Electrogastrography Abnormalities in Pediatric Gastroduodenal Disorders: A Systematic Review and Meta-analysis. J. Pediatr. Gastroenterol. Nutr. 73, 9–16 (2021).

6. Carson, D. A. et al. Abnormalities on Electrogastrography in Nausea and Vomiting Syndromes: A Systematic Review, Meta-Analysis, and Comparison to Other Gastric Disorders. Dig. Dis. Sci. (2021) doi:10.1007/s10620-021-07026-x.

7. Bhat, S. et al. Gastric Dysrhythmia in Gastroesophageal Reflux Disease: A Systematic Review and Meta-Analysis. Esophagus Accepted, (2021).

8. Varghese, C. et al. Clinical associations of functional dyspepsia with gastric dysrhythmia on electrogastrography: A comprehensive systematic review and meta-analysis. Neurogastroenterol. Motil. 33, e14151 (2021).

9. Carson, D. A., O’Grady, G., Du, P., Gharibans, A. A. & Andrews, C. N. Body surface mapping of the stomach: New directions for clinically evaluating gastric electrical activity. Neurogastroenterology & Motility vol. 33 (2021).

10. Calder, S. et al. An automated artifact detection and rejection system for body surface gastric mapping. Neurogastroenterol. Motil. e14421 (2022).

11. Bortolotti, M. Electrogastrography: a seductive promise, only partially kept. The American journal of gastroenterology vol. 93 1791–1794 (1998).

12. Verhagen, M. A., Van Schelven, L. J., Samsom, M. & Smout, A. J. Pitfalls in the analysis of electrogastrographic recordings. Gastroenterology 117, 453–460 (1999).

13. Gharibans, A. A. et al. A novel scalable electrode array and system for non-invasively assessing gastric function using flexible electronics. Neurogastroenterol. Motil. e14418 (2022).

14. Gharibans, A. A., Coleman, T. P., Mousa, H. & Kunkel, D. C. Spatial Patterns From High-Resolution Electrogastrography Correlate With Severity of Symptoms in Patients With Functional Dyspepsia and Gastroparesis. Clin. Gastroenterol. Hepatol. 17, 2668–2677 (2019).

15. Sebaratnam, G. et al. Standardized system and App for continuous patient symptom logging in gastroduodenal disorders: Design, implementation, and validation. Neurogastroenterol. Motil. e14331 (2022).

16. Gharibans, A. A. et al. Gastric dysfunction in patients with chronic nausea and vomiting syndromes defined by a novel non-invasive gastric mapping device. bioRxiv (2022) doi:10.1101/2022.02.07.22270514.

17. Somarajan, S. et al. The effect of chronic nausea on gastric slow wave spatiotemporal dynamics in children. Neurogastroenterol. Motil. 33, e14035 (2021).

18. Chen, J. & McCallum, R. W. Electrogastrography: measuremnt, analysis and prospective applications. Med. Biol. Eng. Comput. 29, 339–350 (1991).

19. Wolpert, N., Rebollo, I. & Tallon-Baudry, C. Electrogastrography for psychophysiological research: Practical considerations, analysis pipeline, and normative data in a large sample. Psychophysiology (2020) doi:10.1111/psyp.13599.

20. Huizinga, J. D., Hussain, A. & Chen, J.-H. Interstitial cells of Cajal and human colon motility in health and disease. Am. J. Physiol. Gastrointest. Liver Physiol. 321, G552–G575 (2021).

21. Dinning, P. G. et al. Quantification of in vivocolonic motor patterns in healthy humans before and after a meal revealed by high-resolution fiber-optic manometry. Neurogastroenterology & Motility vol. 26 1443–1457 (2014).

22. Abell, T. L. et al. Consensus recommendations for gastric emptying scintigraphy: a joint report of the American Neurogastroenterology and Motility Society and the Society of Nuclear Medicine. J. Nucl. Med. Technol. 36, 44–54 (2008).

23. Xu, W. et al. Sa1493: GASTRIC ELECTROPHYSIOLOGICAL ABNORMALITIES DEFINED BY BODY SURFACE GASTRIC MAPPING CORRELATE WITH SYMPTOM BURDEN IN PATIENTS WITH LONG-TERM TYPE 1 DIABETES. Gastroenterology vol. 162 S–396 (2022).

24. Varghese, C. et al. Normative reference range for body surface gastric mapping evaluations of gastric motility using Gastric Alimetry: spectral analysis. In submission doi:DOI to follow.

25. Alighaleh, S., Cheng, L. K., Grady, G. O., Bartlett, A. & Paskaranandavadivel, N. Sa1492: HIGH-RESOLUTION MAPPING OF HUMAN GASTRIC SLOW-WAVE IN RESPONSE TO PACING. Gastroenterology vol. 162 S–396 (2022).

26. O’Grady, G., Gharibans, A. A., Du, P. & Huizinga, J. D. The gastric conduction system in health and disease: a translational review. Am. J. Physiol. Gastrointest. Liver Physiol. 321, G527–G542 (2021).

27. Chang, F.-Y. Electrogastrography: basic knowledge, recording, processing and its clinical applications. J. Gastroenterol. Hepatol. 20, 502–516 (2005).

28. Yin, J. & Chen, J. D. Z. Electrogastrography: methodology, validation and applications. J. Neurogastroenterol. Motil. 19, 5–17 (2013).

29. Simonian, H. P. et al. Multichannel Electrogastrography (EGG) in Normal Subjects: A Multicenter Study. Digestive Diseases and Sciences vol. 49 594–601 (2004).

30. Parkman, H. P., Hasler, W. L., Barnett, J. L. & Eaker, E. Y. American Motility Society Clinical GI Motility Testing Task Force. Electrogastrography: a document prepared by the gastric section of the American Motility Society Clinical GI Motility Testing Task Force. Neurogastroenterol Motil Apr;15(2):89–102, (2003).

31. Koch, K. L. Handbook of Electrogastrography. (Oxford University Press, 2003).

32. O’Grady, G. et al. Origin and propagation of human gastric slow-wave activity defined by high-resolution mapping. American Journal of Physiology-Gastrointestinal and Liver Physiology vol. 299 G585–G592 (2010).

33. Berry, R. Functional physiology of the human terminal antrum defined by high-resolution electrical mapping and computational modeling. Am. J. Physiol. Gastrointest. Liver Physiol 311, G895–G902 (2016).

34. Dinning, P. G. Quantification of in vivo colonic motor patterns in healthy humans before and after a meal revealed by high-resolution fiber-optic manometry. Neurogastroenterol. Motil 26, 1443–1457 (2014).

35. O’Grady, G. et al. Abnormal initiation and conduction of slow-wave activity in gastroparesis, defined by high-resolution electrical mapping. Gastroenterology 143, 589-598.e3 (2012).

36. Angeli, T. R. Loss of Interstitial Cells of Cajal and Patterns of Gastric Dysrhythmia in Patients With Chronic Unexplained Nausea and Vomiting. Gastroenterology 149, 56–66 (2015).

37. Angeli, T. R. et al. Loss of Interstitial Cells of Cajal and Patterns of Gastric Dysrhythmia in Patients With Chronic Unexplained Nausea and Vomiting. Gastroenterology 149, 56-66.e5 (2015).

